# Association between sleep-disordered breathing and moderate to severe depression in systemic lupus erythematosus: the TRUMP^2^-SLE study

**DOI:** 10.1101/2024.08.20.24312299

**Authors:** Yuichi Ishikawa, Nao Oguro, Takanori Ichikawa, Dai Kishida, Natsuki Sakurai, Chiharu Hidekawa, Kenta Shidahara, Keigo Hayashi, Yoshia Miyawaki, Yasuhiro Shimojima, Ryusuke Yoshimi, Ken-ei Sada, Nobuyuki Yajima, Noriaki Kurita

## Abstract

**Objective:** Depression is the most frequent mood disorder that impairs quality of life and medication adherence in patients with systemic lupus erythematosus (SLE). Although sleep-disordered breathing (SDB) is a contributor to depression in the general population, its prevalence in SLE patients and its impact on depression are not clear. We employed a clinical epidemiologic approach to examine them in a multicenter cohort of SLE patients.

**Methods:** This was a cross-sectional study of 414 Japanese adults with SLE at five university hospitals. The main exposure was high-risk SDB, assessed with the Berlin Questionnaire. The main outcome was moderate to severe depression assessed using the Patient Health Questionnaire-9. Poisson regression models were fitted with a robust error variance to estimate adjusted prevalence ratios (aPRs).

**Results:** The mean age was 47.5 years and the mean body mass index (BMI) was 22.1 kg/m^2^. The prevalence of high-risk SDB was 15.2% (95% confidence interval [95% CI] 11.9%–19.0%). The prevalence of moderate or severe depression was 19.1% (95% CI 15.4%–23.2%). High-risk SDB was associated with a greater likelihood of moderate to severe depression (aPR 2.62, 95% CI 1.62–4.24). All the 1-, 2-, and 3-positive risk categories for SDB were associated with moderate to severe depression, in a dose-dependent manner.

**Conclusion:** Among patients with SLE, SDB is common, and high-risk SDB is strongly associated with moderate to severe depression. The signs and symptoms of SDB should prompt a simultaneous evaluation for concomitant depression.

**KEY MESSAGES:**

- SDB and depression are common among SLE patients with relatively low disease activity.
- High-risk SDB was associated with a greater prevalence of moderate to severe depression.
- SDB should be evaluated in SLE patients with concomitant depression.

## INTRODUCTION

Systemic lupus erythematosus (SLE) is a chronic disease characterised by periods of exacerbations and remissions. Patients with SLE are commonly affected by depression (reported prevalence: 8.7%–78.6%). (1,2) Occurrence of depression in SLE patients has been shown to impair their quality of life (QOL) (3,4) and medication adherence. (5,6) Therefore, it is imperative to identify factors contributing to depression and develop strategies to alleviate depression in patients with SLE.

Sleep-disordered breathing (SDB) disorder, including obstructive sleep apnoea (OSA), is a group of disorders characterised by abnormal breathing patterns during sleep. Research has shown that SDB is associated with depression in the general population. (7) Inflammation associated with SLE and airway narrowing associated with glucocorticoid (GC)-induced obesity may increase the risk of OSA and exacerbate SDB. (8) However, the prevalence of SDB among SLE patients and its impact on depression in this population are not well understood.

The reported prevalence of moderate or severe OSA in SLE patients ranges from 21.4 to 23.6%. (8,9) However, most previous studies had small sample sizes, ranging from 14 to 42 patients. Thus, despite the substantial coexistence of sleep disorders among patients with SLE (56.0%–80.5%), there is a lack of robust epidemiologic data on the prevalence of potential SDB in these patients. (10) Although a strong association between sleep disturbances and depression is noted, (11) the impact of SDB, a phenotype of sleep disorders, on depression is also inconsistent due to insufficient sample size and the confounding influence of covariates such as GC use and age. (8,9) From a mechanistic perspective, cytokine imbalances associated with sleep disturbances in SLE may induce a pro-inflammatory state, (12) and the involvement of inflammatory cytokines in the pathophysiology of mood disorders has been postulated. (1) Given this context, a clinical-epidemiological study examining the association between SDB and depression may provide insights into the mechanisms of these complications and help inform preventive measures.

The objective of this study was to examine the prevalence of depression and high-risk SDB patients using a multicenter cohort of patients with SLE and analyse the impact of SDB on depression.

## METHODS

### Study design and settings

This was a cross-sectional study using data from the TRUMP^2^-SLE project which is a multicenter cohort study conducted at five academic medical centres. The data for the present study were collected between March 2021 and March 2023.

### Participants and data collection

The inclusion criteria were as follows: (1) SLE patients diagnosed based on the revised 1997 American College of Rheumatology classification criteria; (13) (2) age ≥20 years; (3) receiving regular care by rheumatologists at the participating center, and (4) having the ability to respond to the questionnaire survey. Patients with dementia or total blindness were excluded from the study. All the enrolled patients were Japanese.

### Exposures

The main exposure was SDB, assessed by the Japanese version of the Berlin Questionnaire (BQ). (14) The BQ consists of three risk categories: snoring (category 1); sleepiness and fatigue during waking time (category 2); obesity (body mass index (BMI) ≥30 kg/m^2^) and hypertension (category 3). The SDB risk was graded by the number of positive categories. Categories 1 and 2 were considered positive if their total respective scores were ≥2. Category 3 was considered positive if the score was ≥1.

Patients with ≥2 positive categories were considered at high risk of SDB, while patients with only 1 or no positive categories were considered low risk. (15) All patients completed the questionnaire either in the waiting room or at home. The questionnaire included assurances that the attending physician would not review the responses and that the responses would only be used at the central facility for data aggregation.

### Outcome

The main outcome was moderate or severe depression, defined as ≥10 points of the Japanese version of the Patient Health Questionnaire (PHQ)-9. (16) The PHQ-9 is a self-reported questionnaire used to identify depression and evaluate its severity. (17,18) The performance of the Japanese version of the PHQ-9 for depression in primary care has been validated; its sensitivity and specificity for major depressive disorder according to the aforementioned cutoffs were 92.5% and 77%, respectively. (16)

### Measurement of covariates

Based on previous studies and clinical insights from rheumatologists, the following variables with demonstrated or plausible association with both SDB and depression were selected as confounding variables: age (19), sex (19,20), BMI, habitual smoking, habitual alcohol consumption, disease activity, daily GC dose, GC pulse, and non-communicable diseases requiring medication such as diabetes, hypertension, dyslipidemia, and hyperlipidemia. Disease activity was measured using the Systemic Lupus Erythematosus Disease Activity Index 2000 (SLEDAI-2K) (21) by the attending physician. Daily GC dose was documented as prednisolone equivalents. Hypertension was considered present if any of the following were prescribed: angiotensin-converting enzyme inhibitors, angiotensin receptor blockers, calcium channel blockers, beta-blockers, anti-aldosterone drugs, diuretics or other anti-hypetensive drugs. Dyslipidemia was considered to be present if any of the following were prescribed: statins, fibrates, cholesterol absorption inhibitors, eicosapentaenoic acid/docosahexaenoic acid, or other hypolipidemic drugs. Diabetes was considered to be present if any of the following were prescribed: DPP4 inhibitors (including GLP-1 agonists), glinides, alpha-glucosidase inhibitors, thiazolidinedione derivatives, sulfonylureas, SGLT-2 inhibitors, biguanides, insulin or other antihyperglycemic drugs.

### Statistical analysis

Continuous variables were expressed as mean ± standard deviation (SD), while categorical variables were expressed as frequency (percentage). The association between high-risk SDB and moderate to severe depression was evaluated using the Poisson regression model with a robust error variance to estimate adjusted prevalence ratios (aPR) adjusted for the abovementioned potential confounders. (22) The rationale for this model is that using odds ratios overestimates the PRs due to the non-rare prevalence of primary outcomes. In addition, the association between the number of risk categories for SDB and moderate to severe depression was also examined. Missing covariates were addressed using multiple imputations. Thirty imputations were performed by multiple imputations with chained equations, assuming that the analysed data were missing at random. (23) *p*_<_0.05 was considered indicative of statistical significance. Statistical analyses were performed using STATA /SE, version 17.0 (StataCorp, College Station, TX, USA) and R version 4.2.2 (R Foundation for Statistical Computing, Vienna, Austria).

## RESULTS

### Participant characteristics and the prevalence of high-risk SDB

A total of 448 patient records were collected. Of these, 34 patients with missing data regarding main exposure and/or the primary outcome were excluded. Finally, 414 patients were included in the analysis (Figure 1). The characteristics of the study population (N = 414) are summarised in Table 1. The prevalence of high-risk SDB was 15.2% (n = 63, 95% confidence interval [95% CI] 11.9%–19.0%). The percentages of patients who had one, two, and three positive risk categories were 35.8% (n = 148), 13.0% (n = 54), and 2.2% (n = 9), respectively. The characteristics of patients in the high-risk SDB and low-risk SDB groups are also summarised in Table 1. Patients in the high-risk SDB group were significantly older (age: 54.0 vs 46.3 y) and had significantly higher BMI (24.9 vs 21.6 kg/m^2^) than those in the low-risk SDB group. In addition, the high-risk SDB group had a lower proportion of women (75.0% vs 89.5%) and a higher prevalence of hypertension (66.0% vs 29.0%) and current smokers (18.2% vs 9.2%).

**Figure 1.**
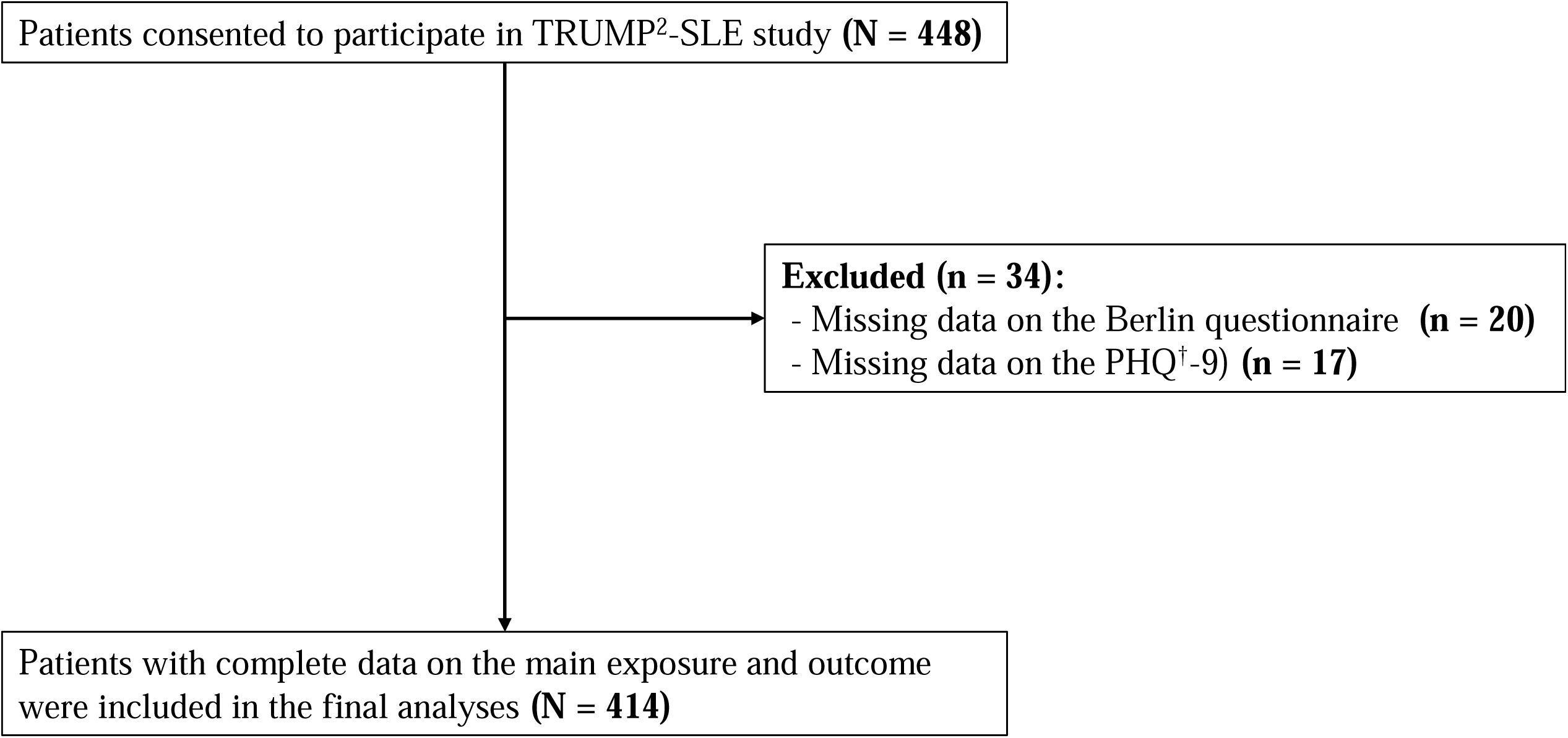
Flowchart of patients.

**Table 1.**
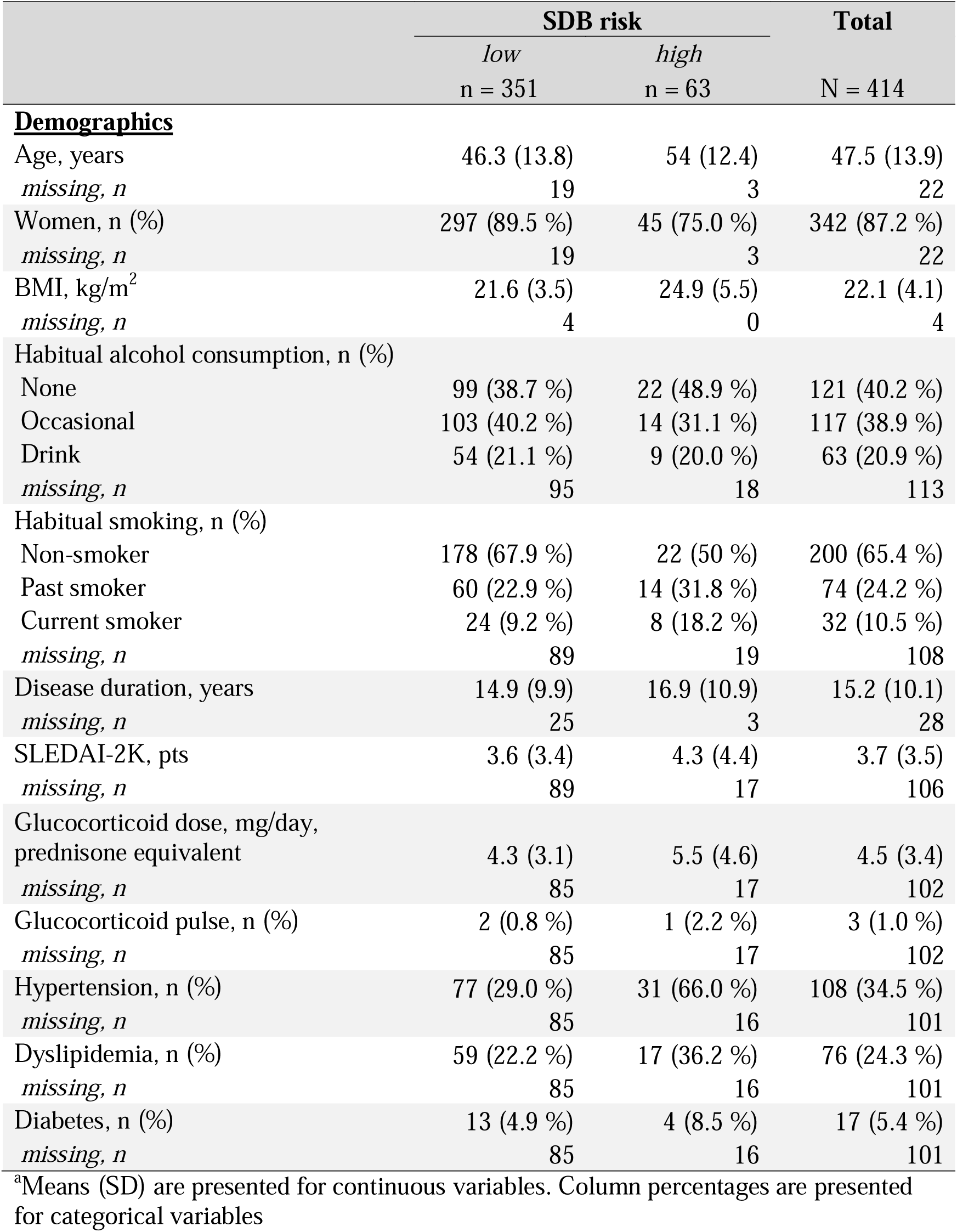
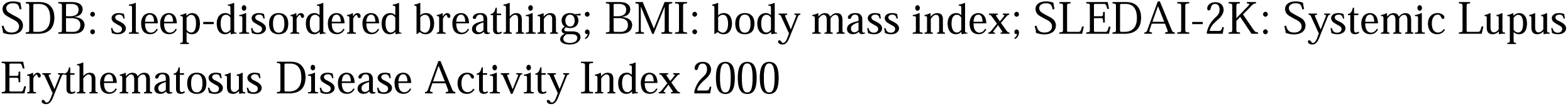
Characteristics of the study population disaggregated by sleep-disordered breathing risk (N=414)

### Prevalence of moderate or more depression

The overall prevalence of moderate to severe depression in this study was 19.1% (n = 79, 95% CI 15.4%–23.2%). The high-risk SDB group had a higher prevalence of moderate or more severe depression (39.7% [n=54] vs 15.4% [n=25]).

### Association between high-risk SDB and moderate or more severe depression

The association of high-risk SDB with moderate to severe depression and covariates is shown in Table 2. High-risk SDB was associated with moderate to severe depression (adjusted prevalence ratio (aPR) 2.62, 95% CI 1.62–4.24). Older age was associated with a lower likelihood of having moderate to severe depression (per 10-year higher: aPR 0.78, 95% CI 0.65–0.93). Compared with those with zero risk category for SDB, those with ≥1 positive risk category was also associated with moderate to severe depression in a dose-dependent manner (Table 3: one category positive, aPR 2.83, 95% CI 1.65–4.85; two categories positive, aPR 4.90, 95% CI 2.54–9.46; three categories positive, aPR 5.12, 95% CI 2.21–11.83).

**Table 2.** The association of moderate or severe depression with high-risk sleep-disordered breathing and covariates.

|  | Adjusted prevalence ratio,<br>point estimate (95% CI) | *P*-value |
| --- | --- | --- |
| **High-risk SDB (vs. Low-risk SDB)** | **2.62 (1.62–4.24)** | **<0.001** |
| **Age, per 10-year increment** | **0.78 (0.65–0.93)** | **0.007** |
| Women (vs Men) | 1.08 (0.59–1.98) | 0.812 |
| BMI, per 1 kg/m<sup>2</sup> increment | 1.02 (0.97–1.06) | 0.478 |
| Habitual alcohol consumption |  |  |
| None | Reference |  |
| Occasional | 0.93 (0.58–1.49) | 0.761 |
| Drink | 0.64 (0.32–1.27) | 0.205 |
| Habitual smoking |  |  |
| Non-smoker | Reference |  |
| Past smoker | 1.44 (0.88–2.37) | 0.151 |
| Current smoker | 1.79 (0.981–3.28) | 0.058 |
| SLEDAI-2K, per 1-point increment | 1.004 (0.95–1.06) | 0.875 |
| Glucocorticoid dose, per 1 mg/day increment | 1.04 (0.98–1.10) | 0.191 |
| Glucocorticoid pulse | 0.95 (0.26–3.53) | 0.940 |
| Hypertension | 1.10 (0.67–1.79) | 0.713 |
| Dyslipidemia | 0.64 (0.35–1.18) | 0.153 |
| Diabetes | 1.14 (0.45–2.88) | 0.784 |

3 The prevalence ratios were estimated by fitting a Poisson regression model with a 4 robust standard variance including all the above variables.
5 Bold font indicates significance at $P < 0.05$ in the *P*-value column. SDB: sleep- 6 disordered breathing; BMI: body mass index; SLEDAI-2K: Systemic Lupus 7 Erythematosus Disease Activity Index 2000

**Table 3.** Table 3. The associations of moderate or severe depression with the number of risk categories for sleep-disordered breathing and covariates.

|  | Adjusted prevalence ratio,<br>point estimate (95% CI) | *P*-value |
| --- | --- | --- |
| The number of risk categories for SDB |  |  |
| None | Reference |  |
| **1-category** | **2.83 (1.65–4.85)** | **<0.001** |
| **2-categories** | **4.90 (2.54–9.46)** | **<0.001** |
| **3-categories** | **5.12 (2.21–11.83)** | **<0.001** |
| **Age, per 10-year increment** | **0.78 (0.65–0.93)** | **0.006** |
| Women (vs Men) | 1.13 (0.61–2.11) | 0.691 |
| BMI, per 1 kg/m<sup>2</sup> increment | 1.01 (0.97–1.05) | 0.532 |
| Habitual alcohol consumption |  |  |
| None | Reference |  |
| Occasional | 0.96 (0.59–1.54) | 0.856 |
| Drink | 0.64 (0.33–1.26) | 0.197 |
| Habitual smoking |  |  |
| Non-smoker | Reference |  |
| Past smoker | 1.45 (0.88–2.40) | 0.149 |
| Current smoker | 1.59 (0.88–2.89) | 0.126 |
| SLEDAI-2K, per 1-point increment | 1.00 (0.95–1.05) | 0.993 |
| Glucocorticoid dose, per 1 mg/day increment | 1.04 (0.98–1.10) | 0.200 |
| Glucocorticoid pulse | 1.06 (0.24–4.62) | 0.934 |
| Hypertension | 1.01 (0.63–1.63) | 0.968 |
| Dyslipidemia | 0.61 (0.33–1.13) | 0.116 |
| Diabetes | 1.19 (0.47–3.00) | 0.706 |

3 The prevalence ratios were estimated by fitting a Poisson regression model with a 4 robust standard variance including all the above variables.
5 Bold font indicates significance at $P < 0.05$ in the *P*-value column. SDB: sleep- 6 disordered breathing; BMI: body mass index; SLEDAI-2K: Systemic Lupus 7 Erythematosus Disease Activity Index 2000

## DISCUSSION

### Summary of the key results

This multicenter study revealed a high prevalence of moderate to severe depression and high-risk SDB (18.9% and 15.4%, respectively) among patients with SLE. In addition, high-risk SDB was associated with a greater prevalence of moderate to severe depression. Furthermore, the prevalence of moderate to severe depression increased with an increase in the number of positive BQ categories in a dose-dependent manner. While several studies have investigated the prevalence of OSA among patients with SLE, to the best of our knowledge, this is the first large-scale epidemiologic study examining the prevalence of SDB in this population. The prevalence rates of moderate to severe OSA in previous studies (range, 21.4–23.6%) were higher than that of high-risk SDB in the present study. (8,9) Furthermore, depression scores assessed by the Becks Depression Inventory have yielded inconsistent results. One study showed that it is not associated with moderate-to-severe OSA (8) while another study reported its association with apnoea/hypopnea index. (9) Nevertheless, the small number of subjects in previous studies as well as the higher BMI and younger age of the subjects compared to the present study may explain the differences in results. Our study provides a more robust finding that SDB can partly explain the already identified correlation between sleep disturbances and depression among patients with SLE. (11)

There are several clinical implications of this study. First, it may be clinically relevant to proactively detect symptoms suggestive of SDB in SLE patients presenting with depression. Indeed, the increased risk of depression associated with even a low risk of SDB in our study supports this notion. Second, SDB may be a modifiable factor that should be considered when exploring non-pharmacologic approaches to address depression among patients with SLE. Non-pharmacologic interventions such as continuous positive airway pressure therapy and oral appliances are available for SDB. (24,25)

There are some strengths of this study. The present study demonstrated an association between SDB risk and depression, including a dose-response relationship, even after adjusting for clinically important potential confounders in a large sample of patients. In addition, the multicenter study design enhances the generalizability of the findings. However, some limitations of this study should be considered while interpreting the results. First, high-risk SDB was classified solely based on the BQ. The BQ has a somewhat low sensitivity and positive predictive value for diagnosing true SDB. (15,26) However, the BQ is a practical instrument for epidemiological studies involving large numbers of subjects, (27,28) because of its convenience, efficiency, and good sensitivity. Second, sex hormones such as oestrogen and progesterone were unmeasured confounders. These hormones are potential risk factors for SDB and are also associated with the development of depression. (29) Third, owing to the cross-sectional nature of the study, the possibility of reverse causality in the relationship between SDB and moderate or severe depression should be considered. For example, atypical depression may lead to increased appetite and weight gain, which may result in high-risk SDB. (30) However, the relationship between high-risk SDB and depression in the present study was independent of BMI. Fourth, this study was conducted in Japan. The prevalence of SDB is reported to vary by race and needs to be verified in non-Asian racial groups. (31)

## CONCLUSIONS

Among patients with SLE with relatively low disease activity, both SDB and moderate to severe depression were common, and high-risk SDB was associated with moderate to severe depression. Our findings highlight the importance of investigating signs and symptoms of SDB as even minor SDB risk is strongly associated with moderate to severe depression.

## ACKNOWLEDGEMENTS

The authors are grateful to all collaborators in the TRUMP^2^-SLE project. The authors also thank a patient organisation, the Rheumatic Disease & Vasculitis Support Network in JAPAN (KOSAPO), a Specified Nonprofit Corporation, and Associate Professor Haruka Nakada of the School of Health Innovation, Kanagawa University of Human Services, for their support in conducting the Patient and Public Involvement.

## AUTHOR CONTRIBUTIONS

All authors fulfil the International Committee of Medical Journal Editors (ICMJE) criteria for authorship. YI, NY, and NK conceptualised and designed the study. All authors made contributions to data collection. YI and NK provided substantial contributions to the analysis and interpretation of data. YI wrote the first draft of the manuscript. All authors contributed to drafting the article and approved the final version of the article for publication. NY and NK supervised the interpretation of data and manuscripts.

## DATA AVAILABILITY STATEMENT

The datasets generated and/or analysed during the study are available from the corresponding author upon reasonable request.

## ETHICS STATEMENTS

### Patient consent for publication

The patient’s informed consent was waived because the study was conducted using anonymous data derived from unmarked questionnaires.

### Patient and Public Involvement

We discussed with KOSAPO how we will disseminate our research results after the article is published. The study results will be communicated through KOSAPO’s website (https://www.rheumatic-disease-community.org/) and social networking sites (X and Instagram). We also consulted with the KOSAPO on the statement to be sent out.

### Ethics approval

This study complied with the Declaration of Helsinki and the Ethical Guidelines for Medical and Biological Research Involving Human Subjects in Japan. The study protocol was approved by the Institutional Ethical Board of Showa University Hospital (approval number: 22-130-B).

